# Leveraging Predictive AI and LLM-Powered Trial Matching to Improve Clinical Trial Recruitment: A Usability Assessment of Trialshub

**DOI:** 10.64898/2026.04.17.26351107

**Authors:** Paa-Kwesi Blankson, Shakira Hussien, Ferdose Idris, Geannene Trevillion, Ali Aslam, Amir Afani, Phenesse Dunlap, Joyline Chopkorir, Paolo Melgarejo, Muhammed Idris

## Abstract

**Background:** Recruitment remains a major barrier to timely clinical trial completion. Trialshub is an LLM-powered, chat-based platform intended to help users identify relevant trials and connect with coordinators to streamline recruitment workflows.

**Objective:** To evaluate the perceived usability and operational value of Trialshub, and identify implementation considerations for real-world deployment.

**Methods:** A usability test was conducted at Morehouse School of Medicine for the Trialshub application. Purposively selected participants included clinical research coordinators and individuals with and without clinical trial search experience. Participants completed a pre-test survey assessing demographics, digital health information behaviors, and familiarity with AI tools, followed by a moderated usability session using a Trialshub prototype. Users completed scenario-based tasks (locating a breast cancer trial, reviewing results, and initiating coordinator contact) using a think-aloud protocol. Task ratings, screen recordings, and transcribed feedback were analyzed descriptively and thematically, and reported.

**Results:** Participants reported high comfort with using digital tools and moderate-to-high familiarity with AI. Trialshub’s chat-first design, guided prompts, and checklist-style eligibility display were perceived as intuitive and reduced cognitive load. Fast access to trials and the coordinator-contact workflow were viewed positively. Key usability issues included uncertainty at step transitions, insufficient cues for selecting results and next actions, and inconsistent system reliability (loading delays, errors, and broken trial detail pages). Participants also noted redundant questioning due to limited conversational memory, requested improved filtering/sorting, and clearer calls-to-action. All participants indicated that Trialshub has strong potential to meaningfully improve clinical trial processes.

**Conclusions:** Trialshub shows promise for improving trial discovery and recruitment workflows, with identified design implications for real-world deployment.

## INTRODUCTION

Clinical trials are foundational to the advancement of medical science, playing a critical role in the development of safe and effective therapies, diagnostics, and medical technologies that improve patient outcomes and enhance overall quality of life.^1^ By rigorously evaluating new interventions under controlled and ethically monitored conditions, clinical trials generate robust evidence that informs regulatory decision-making and guides clinical practice around the world.^2^ However, the success of these trials depends not only on scientific rigor but also on the efficiency and integrity of their operational processes, with patient recruitment being a significant component. Recruitment serves as the gateway to study initiation and determines both the pace and generalizability of research. When recruitment falters, trials experience delays, increased costs, and compromised validity.^3,4^

Despite the existence of well-defined guidelines, persistent barriers continue to challenge both patients and coordinators in the recruitment process. Patients face limited awareness of available studies, often due to inadequate communication from healthcare providers or difficulty navigating complex registries.^5,6^ Eligibility criteria, which may be extensive or difficult to interpret, can inadvertently exclude motivated individuals or dissuade participation. Long-standing mistrust in medical research, particularly among minority communities that have experienced historical ethical violations, further hinders engagement. Furthermore, practical constraints, including transportation difficulties, time demands, and financial burdens, disproportionately affect rural and low-income populations.^7,8^

Coordinators likewise experience systemic barriers that impede efficient recruitment. Fragmented or incomplete patient databases complicate the identification of eligible participants, while the pressures of tight recruitment timelines, competition among studies for the same participant pools, and balancing recruitment responsibilities with broader trial operations contribute to coordinator burnout and diminished trial success. These intertwined challenges underscore the urgent need for innovative, scalable, and culturally responsive recruitment tools that strengthen communication, and support coordinators.^9–11^

Trialshub, an advanced large language model (LLM)–powered conversational platform, is proposed as a solution to persistent barriers in clinical trial recruitment. Developed through collaboration among clinicians, data scientists, healthcare advocates, and patients, Trialshub is designed to modernize and simplify both patient-facing and coordinator-facing processes. The platform supports trial discovery by guiding users through a chat-based interface that provides tailored information, clarifies complex eligibility requirements, and helps individuals identify potentially relevant studies. For clinical research coordinators, Trialshub incorporates real-time analytics, automated workflows, and predictive tools intended to reduce manual workload and improve operational efficiency. Within this context, the study examines a central question: does an LLM-powered conversational tool improve trial discovery and the handoff to coordinators without compromising privacy? By addressing usability, workflow clarity, and trust-related considerations, Trialshub may offer a scalable approach to more equitable, efficient, and patient-centered recruitment.

This study therefore aims to evaluate the perceived effectiveness and operational efficiency of Trialshub, as well as to identify potential challenges and considerations that may arise when implementing the platform within real-world clinical research settings.

## METHODS

### Study Design and Participants

This study employed a usability-testing design grounded in typical clinical trial search scenarios to evaluate the ease of navigating and interacting with the Trialshub platform. A mixed-methods approach was used, incorporating think-aloud protocols, scenario-based tasks, structured surveys, and observational analysis to generate a comprehensive understanding of user experience, operational challenges, and system usability. The study was conducted at the Morehouse School of Medicine (MSM).

Participants included clinical research coordinators from MSM’s Clinical Research Center as well as individuals with prior experience searching for clinical trials and other potential users. A purposive sampling strategy was employed to ensure representation of both expert and non-expert users.

### Description of the Application

Trialshub is a chat-based, intuitive platform designed to assist users in identifying suitable clinical trials by interacting with it as they would with a conversational chatbot (Figures 1-3). As users provide information, the system generates tailored guidance and recommendations. When appropriate, Trialshub connects users directly to clinical research coordinators for further communication and scheduling of next steps.

**Figure 1.**
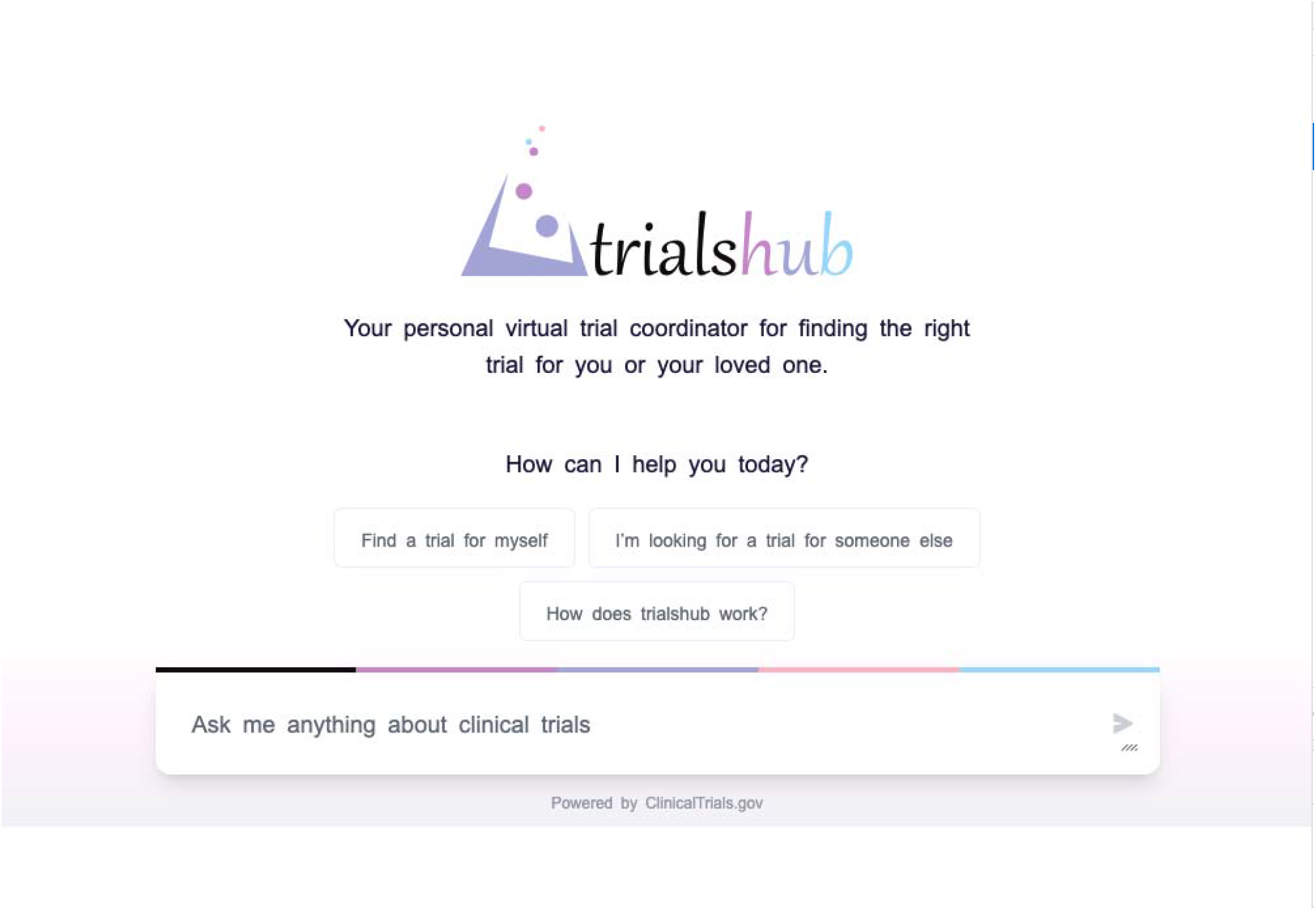
Interface of the Trialshub application home screen

**Figure 2.**
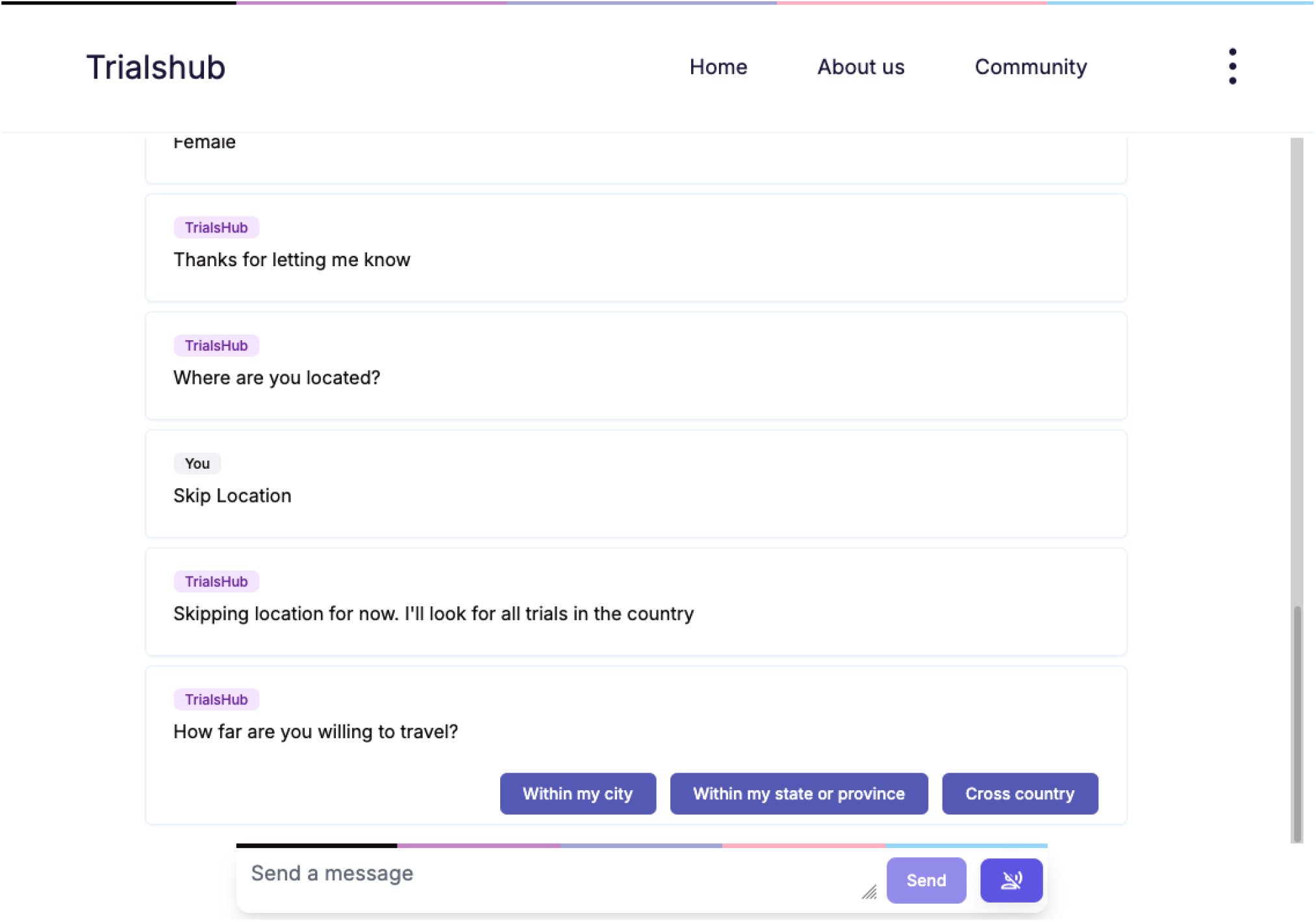
Screenshot of the application’s conversation interface.

**Figure 3.**
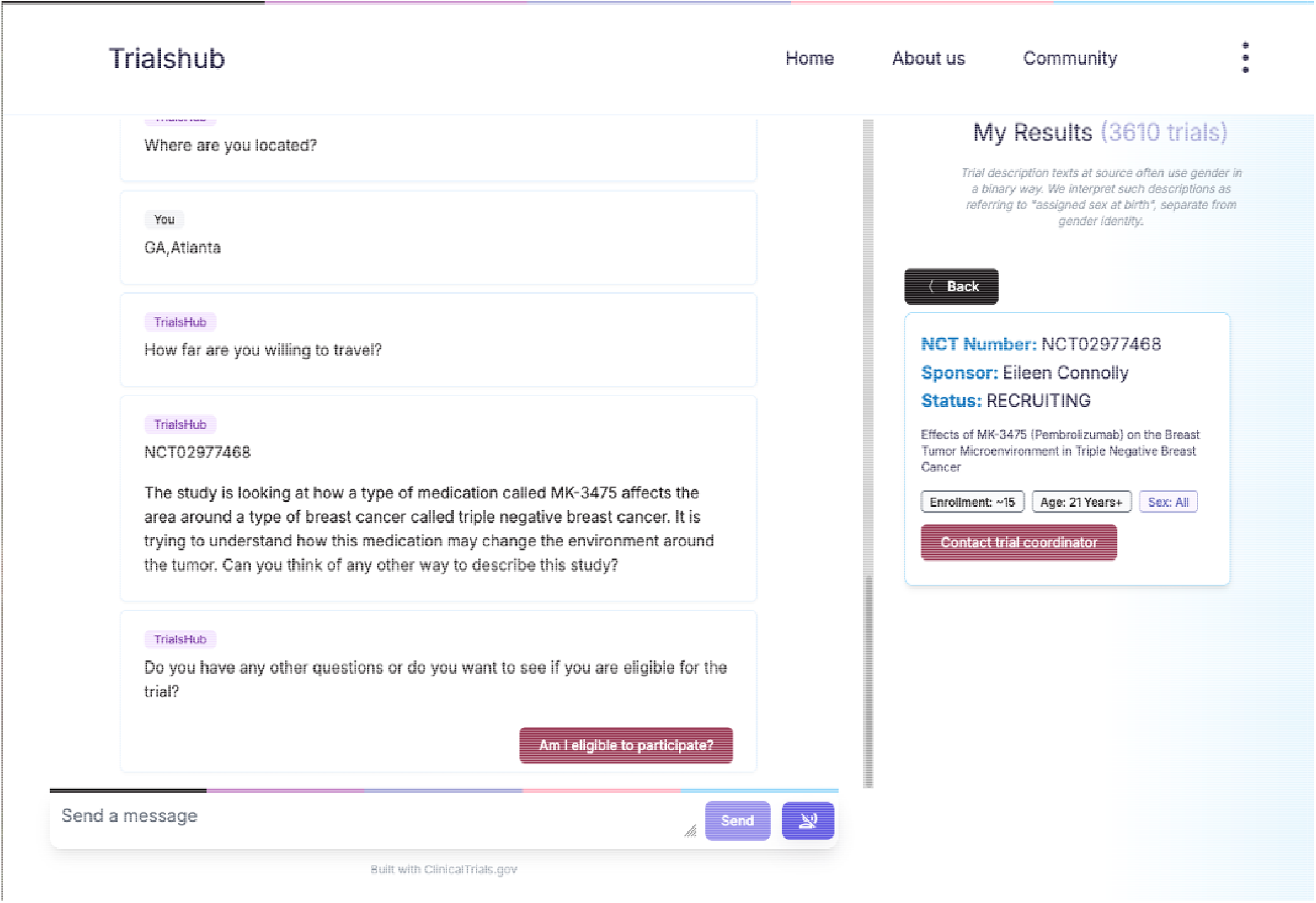
Interface of a trial page in the Trialshub application

### Data Collection Procedures

After providing informed consent, participants completed a structured questionnaire capturing demographic characteristics and background information. Survey items included sociodemographic data; personal or indirect experience with clinical trials; primary device use; frequency of seeking health information online; trusted health-information sources; comfort with adopting new digital tools; and familiarity with AI technologies such as chatbots and predictive systems. Additional questions assessed skills and confidence in accessing and evaluating health information using AI-driven platforms (Appendix A).

Participants were then asked to interact with a prototype version of the Trialshub application and complete a series of predefined tasks (locating a clinical trial related to breast cancer). They were instructed to think aloud while performing each task, verbalizing their actions, expectations, and any confusion or challenges encountered. After each task, participants rated their experience on a 5-piont scale ranging from “extremely easy” to “extremely difficult” and provided additional qualitative feedback. All interactions were video-recorded, screen-captured, and subsequently transcribed.

### Data Analysis

Quantitative data from demographic questionnaires and task ratings were analyzed descriptively. Qualitative data, including think-aloud transcripts, open-ended survey responses, and observational field notes, underwent thematic analysis using iterative coding to identify recurring patterns related to usability challenges, navigation issues, trust, accessibility barriers, intuitiveness of design, and desired features. Comparative analysis was conducted to explore similarities and differences in usability needs between coordinators and general users. Findings from qualitative and quantitative analyses were integrated to develop a comprehensive usability profile of the Trialshub platform. Responses to Likert-scale items were coded numerically from 1 to 5, and summary statistics were calculated as means with corresponding standard deviations.

### Quality Assurance

All data collection instruments, including surveys, observation templates, and usability tasks, were pilot-tested for clarity and relevance. Methodological triangulation combining questionnaire data, usability metrics, and qualitative user feedback was employed to strengthen the validity and consistency of findings. A diverse participant sample was intentionally selected to capture a wide range of user perspectives. Detailed documentation of all procedures was maintained to support replicability and transparency.

### Ethical Considerations

Ethical approval for the study was obtained from the Morehouse School of Medicine Institutional Review Board (IRB #2283200-1). Written informed consent was obtained from all participants, and confidentiality was maintained throughout the study in accordance with institutional and federal guidelines.

## RESULTS

Overall, respondents were mostly young adults and represented a diverse mix of racial and ethnic backgrounds, with the largest proportion identifying as Black or African American (Table 1). Educational attainment was generally high, with most reporting at least a bachelor’s degree. Many participants had prior exposure to clinical trials through searching or considering enrollment, while fewer reported direct participation. Most had not previously used a dedicated trial-finder tool such as ClinicalTrials.gov. Digital engagement was strong: smartphones were the dominant access device, and participants reported regular use of online sources for health information, relying most heavily on clinicians and reputable institutional websites. Participants also reported high comfort adopting new digital tools, moderate-to-high familiarity with AI technologies, and routine use of AI platforms in daily life (Table 1).

**Table 1:** Participant Characteristics and Baseline Digital Health/AI Use (Pre-Test Survey)

| Characteristic | Category | n (%) / Mean (SD) |
| --- | --- | --- |
| Age group | 18–24 | 3 (23.1%) |
|  | 25–34 | 8 (61.5%) |
|  | 35–44 | 2 (15.4%) |
| Gender | Female | 7 (53.8%) |
|  | Male | 5 (38.5%) |
|  | Prefer not to say | 1 (7.7%) |
| Race/ethnicity | Black or African American | 6 (46.2%) |
|  | Black/Arab | 2 (15.4%) |
|  | White | 2 (15.4%) |
|  | Hispanic or Latino | 1 (7.7%) |
|  | Arab | 1 (7.7%) |
|  | Missing | 1 (7.7%) |
| Highest education completed | High school diploma/GED | 1 (7.7%) |
|  | Some college | 2 (15.4%) |
|  | Bachelor's degree | 5 (38.5%) |
|  | Graduate/professional degree | 5 (38.5%) |
| Ever searched/considered joining a trial (self or someone close) | Yes | 8 (61.5%) |
|  | No | 5 (38.5%) |
| Ever participated in a trial (self or someone known) | Yes | 5 (38.5%) |
|  | No | 8 (61.5%) |
| Used ClinicalTrials.gov or another trial-finder tool | Yes | 4 (30.8%) |
|  | No | 9 (69.2%) |
| Primary device | Smartphone | 10 (76.9%) |
|  | Laptop/Desktop | 2 (15.4%) |
|  | Tablet | 1 (7.7%) |
| Frequency of looking up health info online | Very often (daily/almost daily) | 3 (23.1%) |
|  | Often (weekly) | 2 (15.4%) |
|  | Sometimes (monthly) | 5 (38.5%) |
|  | Rarely (few times/year) | 3 (23.1%) |
| Trusted sources of health information (*multi-select*) | Doctor/healthcare provider | 10 (76.9%) |
|  | Government websites | 7 (53.8%) |
|  | Hospital/health system websites | 6 (46.2%) |
|  | Friends/family | 4 (30.8%) |
|  | News media | 3 (23.1%) |
|  | Social media | 1 (7.7%) |
| Comfort using new apps/websites (1–5) | — | 4.54 (0.78) |
| Familiarity with AI technologies (1–5) | — | 4.23 (0.73) |
| AI platform use frequency | Daily | 4 (30.8%) |
|  | Weekly | 5 (38.5%) |
|  | Occasionally | 4 (30.8%) |

**Table 2:**
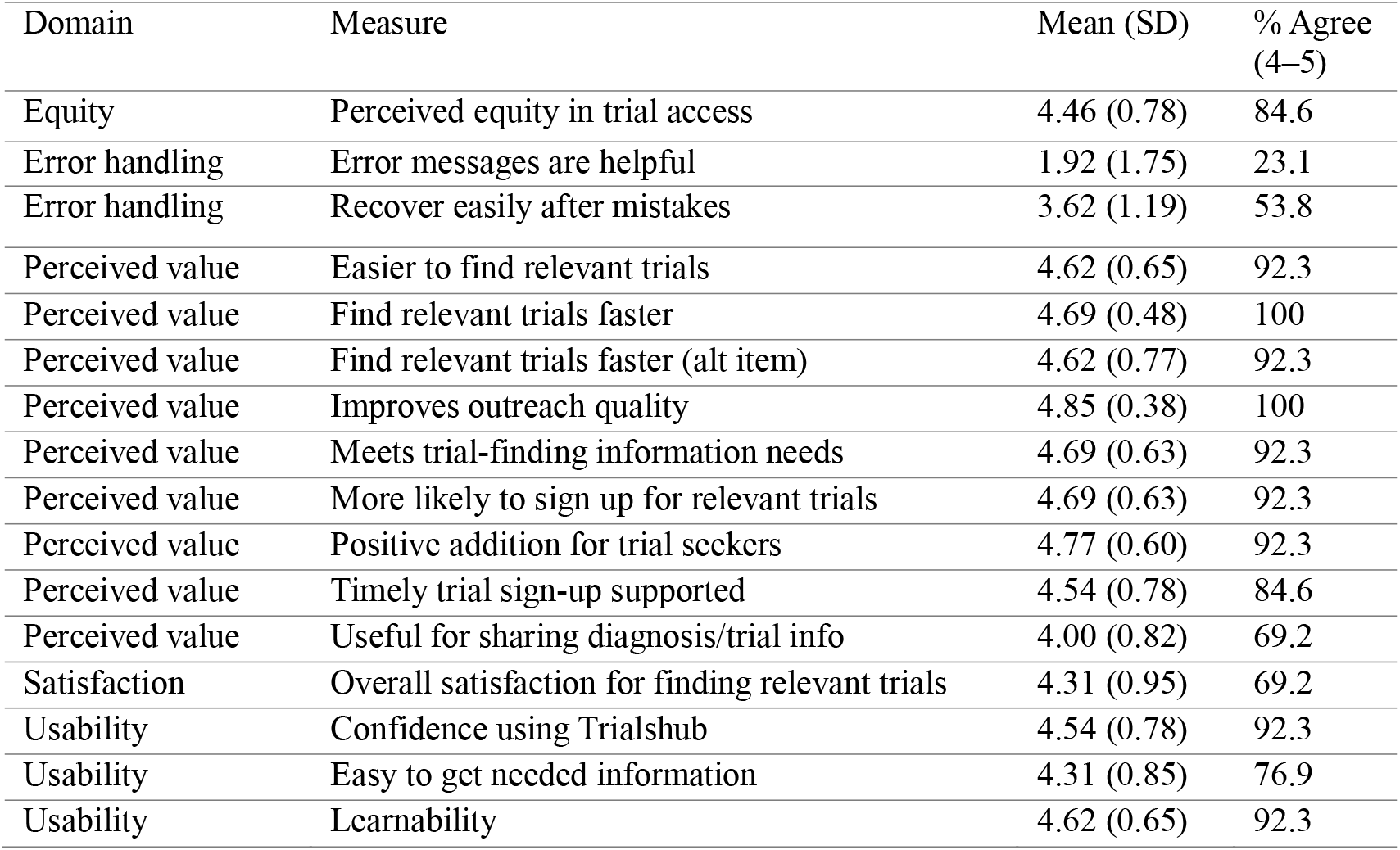
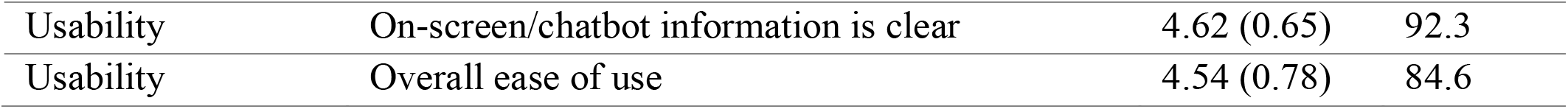
Task Completion Ratings and Perceived Difficulty During the Trialshub Usability Test.

Analysis revealed five major domains: (1) strengths of the current user experience, (2) points of uncertainty and confusion during task completion, (3) insights into user reasoning and interaction patterns, (4) core usability and system reliability pain points, and (5) user-driven feature requests for future iterations.

### Positive user experience and perceived value

Overall, participants described the chat-first interaction as intuitive and approachable, noting that guided prompts reduced cognitive load, particularly at key decision points. Users responded favorably to structured, step-by-step guidance and emphasized the value of prompt bars and suggested inputs, which helped them proceed confidently through tasks. Participants also highlighted the importance of clear error and status messages and appreciated the eligibility-check format, particularly when criteria were presented in a structured checklist style that supported quick scanning and comprehension. The interface aesthetics were viewed positively; the dashboard was described as visually appealing and not overly crowded, reinforcing that the core experience was enjoyable when system performance was stable. Participants valued quick access to trial results and emphasized that speed and responsiveness enhanced perceived usefulness. The process of contacting a clinical coordinator was consistently described as clear and simple, with participants noting that response time was exceptionally fast and the workflow felt efficient. Users also expressed satisfaction with the follow-up flow once an appropriate trial was selected, and they endorsed features such as auto-fill, tags (e.g., age/sex/enrollment indicators), and button-based quick replies as helpful for the interaction.

### Uncertainty, workflow ambiguity, and trust-related concerns

Despite the overall positive impressions, participants encountered points of confusion related to navigation and task flow. Some users requested clearer visual emphasis for key interface elements, including “My Results,” a visible “Selected Trial” header, and explicit cues indicating that results were clickable and that a trial must be selected to view details. Participants also indicated that the pathway from eligibility determination to contacting a coordinator required clearer signposting, including a more prominent call-to-action (CTA) to guide the next step.

Eligibility questions were generally understandable, but participants expressed difficulty when the requested information was not readily known (e.g., gene mutation status), noting that Trialshub should explicitly accommodate uncertainty and provide clear guidance for “unknown” responses. Users also requested clearer labeling of input fields, particularly location, and better guidance on how to resolve loading issues or refresh the page when the interface stalled. Trust and privacy considerations surfaced, with some participants requesting clearer explanations of what information would be shared with coordinators and how it would be used. Some suggested that including coordinator photos or profiles could strengthen credibility and reduce hesitation when transitioning from AI interaction to human contact. Participants also indicated a preference for confirmation feedback (e.g., “Your request was submitted”) and more visible loading indicators to increase confidence that actions were completed successfully.

### User interaction patterns and cognitive load observations

Observational insights suggested that users’ attention was often “captured” by the chat panel, contributing to reduced awareness of the results pane on the right. Participants sometimes experienced prompt paralysis at the start of tasks, reporting uncertainty about what to type and what the system expected. Users also commented on the structure and organization of displayed trial lists, noting that results sometimes lacked intuitive ordering (e.g., non-alphabetical locations). In some cases, participants were uncertain whether they had completed specific tasks correctly, indicating that the system’s completion states and success cues were not sufficiently explicit.

### Critical usability pain points and system reliability issues

Participants reported usability issues related to system performance. Perpetual loading, slow responses, and intermittent errors were described as the most challenging aspects of the experience. Keyboard-based search behaviors were inconsistent, with some users reporting that search input disappeared or returned no results. Trial detail pages were described as unreliable or broken at times, which interrupted task completion. Other issues were associated with conversational memory and context persistence: users noted repeated questions, conversational loops, and a lack of continuity across tasks (e.g., being asked for age after having already provided it). Participants indicated that Trialshub should better retain prior inputs, reduce redundancy, and more clearly communicate when task flows build on earlier interactions. Additional interface concerns included limited ability to handle longer or more conversational free-text queries, and constraints that required users to follow narrow and non-obvious phrasing pathways to proceed.

### Feature requests and recommendations for future development

Participants proposed a range of enhancements aimed at improving efficiency, clarity, and accessibility. A dominant theme was the need for stronger filtering and sorting functionality within the results pane, including the ability to filter by therapy type, keywords, enrollment status, age, sex, proximity/distance, and top matches. Users recommended clearer separation between chat and results panels through color, layout, and visual hierarchy improvements, as well as more familiar messaging conventions (e.g., aligning user messages and system responses on opposite sides). Several participants suggested using pre-specified response options (e.g., yes/no/other) where appropriate. Additional requests included a persistent “New Search” or “Reset chat” control, expandable results and trial detail panes, resizable text input for longer messages, and multiple coordinator contact modalities (call, text, email). Finally, participants expressed interest in secure upload capabilities for documents (e.g., lab results or scans), while noting that such features should remain hidden or clearly labeled until fully implemented to avoid confusion.

Across respondents, Trialshub was generally perceived as valuable and easy to use, particularly for speed of trial discovery and the sense that it could strengthen outreach. Task ratings suggested that most workflows were manageable with low effort, though one task stood out as more effortful, less smooth than the others, consistent with open-ended feedback describing moments of navigation uncertainty and “what to do next” ambiguity. Overall, most respondents indicated the tool met expectations fully or in part, and qualitative responses emphasized that guided prompts and coordinator contact felt particularly helpful when the system was functioning smoothly.

Overall, 92.3% of participants reported that the app met their expectations either fully or partially, indicating broad alignment between users’ needs and the platform’s intended functionality.

## DISCUSSION

This usability evaluation suggests that Trialshub is a promising, user-centered tool for supporting clinical trial discovery and recruitment by combining a chat-first interface, guided prompts, and streamlined pathways to connect users with clinical research coordinators. Overall perceptions were strongly positive, as most participants indicated that the platform met expectations fully or in part, and post-test ratings reflected high perceived value for finding trials faster, improving outreach quality, and supporting trial seekers in identifying relevant opportunities. These findings align with the premise that recruitment success depends heavily on clear, accessible information exchange between potential participants and research teams, and that digital tools can reduce friction in the process by improving navigation, comprehension, and actionability of trial options.^12,13^

Participants consistently endorsed Trialshub’s guided prompts as reducing cognitive load and enabling progress through key decision points. This is particularly relevant in clinical trial recruitment, where potential participants often face complex information, unfamiliar terminology, and eligibility uncertainty.^10,14^ In Trialshub, checklist-style eligibility presentation, button-based quick replies, and structured step-by-step flows were perceived as supportive and intuitive, reinforcing a design principle that recruitment tools should minimize effort at the point of decision. Users also described the dashboard as visually appealing and not crowded, and valued fast access to trial results, factors that can improve engagement and reduce abandonment.

A particularly notable finding was the strong user perception that contacting a coordinator was easy and clear, with exceptionally quick response times. From a recruitment standpoint, this “handoff” from self-service trial exploration to human interaction is critical. Coordinators are often central to building trust, clarifying eligibility, and addressing practical barriers such as transportation, scheduling, and study burden.^15–17^ Trialshub’s ability to facilitate this transition while preserving user momentum represents a meaningful advantage over static trial registries and may reduce drop-off between trial identification and enrollment initiation.

Despite favorable impressions, the usability test identified challenges that could directly affect real-world recruitment outcomes. Participants requested clearer messaging distinguishing search modes, improved labeling for key fields, and prompts that normalize uncertainty, while still providing actionable paths forward. These findings underscore that usability is not only about aesthetics or navigation, but also about supporting real clinical contexts where users may lack technical knowledge, medical documentation, or confidence to answer eligibility questions.

Recruitment remains one of the most persistent threats to clinical trial success, frequently contributing to delayed timelines, increased costs, and underpowered studies. Barriers such as limited awareness, complex eligibility criteria, and logistical constraints are widely recognized, and these issues are amplified for underrepresented populations.^10^ Trialshub directly targets several of these barriers by simplifying trial discovery, lowering the literacy burden through conversational interaction, and connecting users quickly to coordinators who can provide human guidance. Participants’ strong endorsement of Trialshub’s speed, ease of use, and outreach potential suggests the platform may contribute to improved recruitment efficiency and, potentially, improved representativeness of enrolled cohorts, particularly if paired with culturally responsive content and trusted institutional partnerships.

From a public health perspective, improving trial recruitment and retention has downstream benefits for the speed and quality of therapeutic innovation and for the equitable distribution of research benefits.^18^ Underrepresentation in clinical trials limits the generalizability of evidence and can worsen health disparities when treatments are developed and validated without adequate representation of populations most affected by disease.^19,20^ A tool like Trialshub has potential to support more equitable recruitment by improving access to information, lowering barriers to trial discovery, and enabling more efficient navigation for users who might otherwise be excluded due to limited time, limited health literacy, or lack of familiarity with trial registries.

However, achieving this equity promise requires careful attention to trust-building and accessibility. Findings from this study indicate that users want greater transparency and reassurance regarding data sharing and privacy. In communities with heightened mistrust, these elements may be decisive. Additionally, the presence of at least one participant using assistive technology in the pre-test survey underscores the importance of accessibility features (clear scroll cues, readable layouts, adjustable text input, simplified guided modes) to ensure inclusivity across ability levels and device contexts.

Based on the findings, several targeted improvements could strengthen Trialshub’s usability and overall recruitment impact. System reliability should be prioritized by improving loading stability, ensuring trial detail pages consistently render as expected, alongside providing clear recovery guidance when issues occur. Workflow clarity should also be enhanced through explicit, visually prominent calls-to-action at key transition points, particularly when users move from viewing results to selecting a trial, and from eligibility determination to contacting a coordinator. In addition, improving conversational memory is essential to reduce user frustration. Also, the application should explicitly support common uncertainty scenarios and incorporate plain-language tooltips or brief explanations for complex eligibility terms to reduce confusion and prevent dead ends.

This study has limitations. As a usability evaluation, findings are shaped by a modest sample size and may not generalize to all user groups, particularly individuals with lower digital literacy, limited English proficiency, or limited access to smartphones and stable internet. Additionally, participants tested a prototype; perceptions may evolve as technical stability improves and features mature. Future work should include iterative testing across more diverse community settings, and possibly, evaluation of recruitment outcomes such as conversion rates from search to coordinator contact, enrollment initiation, and retention. Assessing effectiveness in real recruitment workflows, especially for underrepresented groups, will be critical to validating Trialshub’s public health value.

## CONCLUSION

Trialshub demonstrates strong potential to streamline clinical trial discovery and recruitment by supporting trial seekers and reducing coordinator burden. By lowering informational and navigational barriers to participation, the platform may help broaden access to clinical research opportunities, particularly for populations historically underrepresented in biomedical research, thereby strengthening the inclusiveness and real-world applicability of trial evidence.

## Data Availability

All data produced in the present study are available upon reasonable request to the authors

